# COVID-19 Utilization and Resource Visualization Engine (CURVE) to Forecast In-Hospital Resources

**DOI:** 10.1101/2020.05.01.20087973

**Authors:** Shih-Hsiung Chou, James T Kearns, Philip Turk, Marc A. Kowalkowski, Jason Roberge, Jennifer S. Priem, Yhenneko J. Taylor, Ryan Burns, Pooja Palmer, Andrew D. McWilliams

## Abstract

**Background:** The emergence of COVID-19 has created an urgent threat to public health worldwide. With rapidly evolving demands on healthcare resources, it is imperative that healthcare systems have the ability to access real-time local data to predict, plan, and effectively manage resources.

**Objective:** To develop an interactive COVID-19 Utilization and Resource Visualization Engine (CURVE) as a data visualization tool to inform decision making and guide a large health system’s proactive pandemic response.

**Methods:** We designed and implemented CURVE using R Shiny to display real-time parameters of healthcare utilization at Atrium Health with projections based upon locally derived models for the COVID-19 pandemic. We used the CURVE app to compare predictions from two of our models –one created before and one after the statewide stay-at-home and social distancing orders (denoted before- and after-SAH-order model). We established parameter settings for best-, moderate-, and worst-case scenarios for pandemic spread and resource use, leveraging two locally developed forecasting models to determine peak date trajectory, resource use, and root mean square error (RMSE) between observed and predicted results.

**Results:** CURVE predicts and monitors utilization of hospital beds, ICU beds, and number of ventilators in the context of up-to-date local resources and provides Atrium Health leadership with timely, actionable insights to guide decision-making during the COVID-19 pandemic. The after-SAH-order model demonstrated the lowest RMSE in total bed, ICU bed, and patients on ventilators.

**Conclusions:** CURVE provides a powerful, interactive interface that provides locally relevant, dynamic, timely information to guide health system decision making and pandemic preparedness.

## Background

The coronavirus disease 2019 (COVID-19) has resulted in a global pandemic of acute respiratory illness.^1–3^ As of April 27, 2020, there were 2,954,222 COVID-19 confirmed cases and 202,597 deaths reported worldwide.^3^ In the United States (US) alone, COVID-19 has spread to all 55 jurisdictions with 957,875 total confirmed cases, 128,673 hospitalizations, and 56,386 deaths.^4,5^ As COVID-19 continues to spread and case numbers rise, health systems have strained to keep pace with demands for hospital and critical care services, challenging hospitals to provide high-quality patient care while managing potentially scarce resources.^6^ Given these concerns, it is imperative that healthcare systems are able to predict increased utilization and proactively plan effective resource allocation, including when and how much to expand access to hospital beds, intensive care unit (ICU) beds, and ventilators.^7,8^

Many studies have reported on the basic susceptible-infected-recovered (SIR) model to forecast how potential COVID-19 cases might impact the capacity of healthcare systems.^9–19^ However, these studies have several limitations: models using national and international infection projections to provide insights for other countries, states, or regions do not: (a) take into consideration variation in transmission and infection rates based on local population characteristics and spatial differences; (b) apply control measures for the type and timing of medical and behavioral interventions including social distancing, border control, and stay at home policies; (c) account for local community resource availability; or (d) adjust for the ability of regional hospital systems to transfer resources between facilities and repurpose spaces for COVID use. Additionally, despite their widespread application in the COVID-19 pandemic, basic SIR models were not developed to forecast hospitalizations, resulting in uncertainty about their accuracy.^19^

In response to the need for actionable data insights, investigators from the Center for Outcomes Research and Evaluation (CORE) at Atrium Health in Charlotte, North Carolina developed a web-based application, or ‘app’, that embeds COVID-19 forecasting models, developed using local data, to guide the health system’s proactive pandemic response. The COVID-19 Utilization and Resource Visualization Engine (CURVE) app helps system leaders have access to the forecast predictions and observed utilization, alongside existing and surge capacity. Here we present our decision support app as an example for other healthcare systems as COVID-19 pandemic planning efforts are likely to continue for the foreseeable future.

## Methods

### Study context and setting

Atrium Health is a large, vertically integrated, not-for-profit healthcare system with over 50 hospitals and 900 care locations in North Carolina, South Carolina, and Georgia. The health system is headquartered in Charlotte, North Carolina, the largest metropolitan region in the Carolinas. In response to the COVID-19 pandemic, Atrium Health activated its Corporate Incident Command structure on March 6, three days after the first case of COVID-19 was reported in North Carolina and five days before Atrium Health diagnosed its first case of COVID-19 on March 11. The primary goal of the Corporate Incident Command is to coordinate resource planning, preparedness, and decision making, while maintaining regular clinical operations and protecting patients and staff.

The development and deployment of the CURVE app was designed for the greater Charlotte region that includes Anson, Cabarrus, Catawba, Cleveland, Gaston, Iredell, Lincoln, Mecklenburg, Rowan, Stanly, and Union counties. The app creation was divided into two phases. The first phase required the development of COVID-19 infection forecast modeling.^20^ The second phase involved user interface construction, emphasizing information accessibility to health system leaders, and configuration with the host server.

We collected bed use information from Atrium Health’s electronic healthcare record (EHR). Unique occupied beds were defined as an occupied bed in a calendar day regardless of the number of patients that used the bed during the day. Unique beds were derived from the hospital, nursing location, room number, and bed number. No restrictions were placed (e.g., department type, observation encounter) on the calculation of total occupied beds per day. A project management model was initiated to ensure rapid execution and to prioritize user requests for CURVE app development and modifications. Stakeholders included Atrium Health’s executive leadership committee, Information and Analytics Services leaders, and the Strategic Services Group. Obtaining stakeholder feedback resulted in a focus on projections for the following domains: hospital utilization, bed capacity, ICU capacity, and ventilator capacity.

### Phase I – forecasting model development

#### Susceptible-Infected-Recovered-Social Distancing-Detection Rate (SIR-D2) model

The COVID-19 infection forecasting model used in the app was an internally developed epidemic SIR model that incorporated social distancing and detection rate (called SIR-D2), where S is the number of individuals that are susceptible to infection in the population, I is the number of individuals that are infected, and R is the number of individuals that are removed from the population via recovery and subsequent immunity or death from infection. The D2 aspect of the model accounts for the stay-at-home intervention implemented in North Carolina and imperfect detection of COVID-19 cases to estimate latent prevalence. The SIR-D2 model was fit to prevalence data from the greater Charlotte region, representing the health system’s primary Charlotte market area. The model is more fully detailed by Turk et al.^20^

#### Determining hospitalization rate

In order to forecast hospital resource demand, we needed to first determine rates of hospitalization. The hospital rate *p* is the rate per person per day at which COVID-19 positive persons transition from an infected state to an implied hospitalized state. We conjecture the hospitalized state mediates the transition from the infected to the removed state in an SIR-D2 model. If we assume an average illness duration of 14 days,^21^ then by a simple binomial argument, we can solve for the proportion of cases requiring hospitalization. Conversely, for a given proportion of cases requiring hospitalization specified in the app, again assuming an average illness duration of 14 days, we solve for the rate. This rate is then multiplied by the prevalence of COVID-19 positive people at time *t*, to give us the incidence of hospitalization at time *t*. The equation to calculate the hospital rate *p* is as follows:

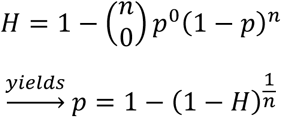

where *H* is the proportion of cases requiring hospitalization, which we estimated from Atrium Health hospital admission data, and *n* is the average illness duration of 14 days described by the CDC.^21^

### Phase II – user interface construction and app development

#### CURVE App Development

We developed the interactive CURVE app using R Shiny (RStudio Inc., Boston, MA).^22^ The inputs default to values determined to be the system standard by weekly stakeholder review. Most input parameters in the app are adjustable by users. The main inputs that drive the CURVE app are as follows: 1) the percentage of total projected infections that would require hospitalization; 2) the proportion of total hospital admissions that would require ICU care; 3) the proportion of patients who are admitted to the ICU who will require ventilator support; 4) the average length of stay in the hospital; 5) the average length of stay in the ICU; 6) the average length of ventilator support when required; and 7) the Atrium Health market share within the grater Charlotte area.

We set up three scenarios (best-, moderate-, and worst-case scenario) with different settings to show a range of potential impacts on the hospital system, due to the uncertain nature of the COVID-19 pandemic. Table 1 gives the parameter settings for each scenario. For planning purposes, our health system chose to use the moderate-case scenario. We adjusted hospitalization, ICU, and on-ventilator percentages for each scenario based on either estimations from the literature or local data trends once we accumulated enough patients.^18,19,23^ Dynamically changing input parameters cause results to cascade through the following charts: 1) hospital census by day shows the projected number of infected patients in the hospital adjusted by the average length of hospital stay; 2) hospital census against capacity by day, including additional surge beds, to indicate if forecasted hospitalization requirements will exceed the healthcare system’s capacities; 3) the observed ICU proportion of hospitalization and observed mechanical ventilator occupancy status with 95% confidence intervals; 4) new daily admissions; 5) the observed hospital census against the projected hospital census; and 6) the predicted actual prevalence.

**Table 1.**
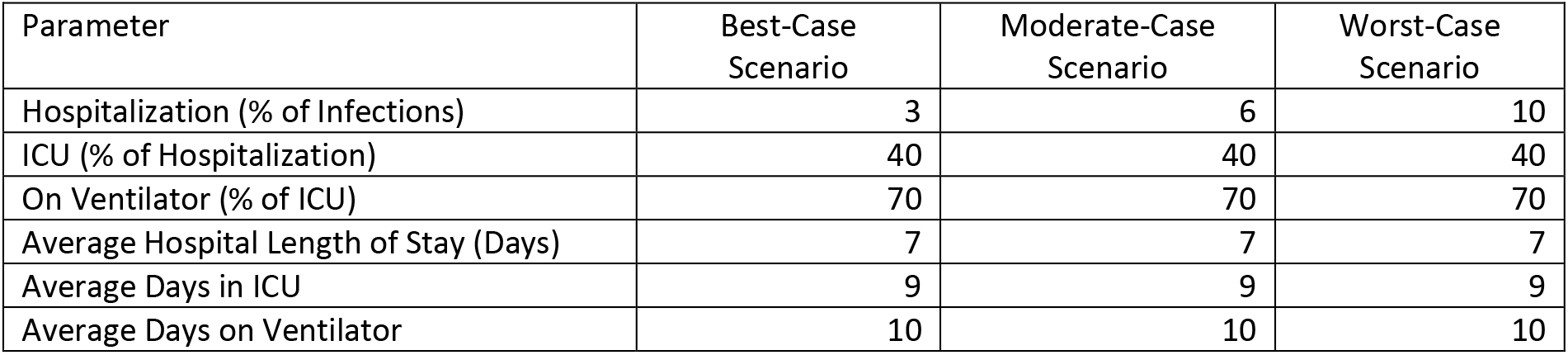
Parameter settings across different scenarios.

| Parameter | Best-Case Scenario | Moderate-Case Scenario | Worst-Case Scenario |
| --- | --- | --- | --- |
| Hospitalization (% of Infections) | 3 | 6 | 10 |
| ICU (% of Hospitalization) | 40 | 40 | 40 |
| On Ventilator (% of ICU) | 70 | 70 | 70 |
| Average Hospital Length of Stay (Days) | 7 | 7 | 7 |
| Average Days in ICU | 9 | 9 | 9 |
| Average Days on Ventilator | 10 | 10 | 10 |

#### Model comparison

We then used the CURVE app to compare predictions from two of our models-one created before and one after the statewide stay-at-home and social distancing orders issued on March 30, 2020 (denoted before-SAH-order and after-SAH-order models hereafter).^24^ Note that the before-SAH-order model was driven by conventional SIR model, which was not adjusted for social distancing and stay-at-home order, i.e., the probability of transmission and exposure rate were not adjusted. We compared the two models’ performance in terms of root mean square error (RMSE) between observed values and predicted results. Finally, using the after-SAH-order model we present all results for best-, moderate-, and worst-case scenario parameter settings including hospital census in all-bed, ICU-bed, ventilator usage, and RMSE (observed vs. predicted).

## Results

The schematic of the proposed R Shiny app reactivity diagram is shown in Figure 1. The app contains four major parts– mod_fun.R, global.R, ui.R, and server.R files. We used the global.R file as an initiation placeholder for importing and wrangling data. We imported data, such as SIR-D2 forecasting simulation results (e.g. infected latent prevalence), default settings, parameters, current bed and ventilator counts, surge beds and additional ventilators, days for preparing additional resources, etc. stored in a csv file for users to easily modify without hard coding in the app. Then global.R feeds the default values and parameters to the input components, followed by calling mod_fun.R to calculate the forecasting numbers, such as initial hospital census, incidence, and new admissions. The generated initial dataset is pushed into a main data hub as a reactiveValues object that can notify any reactive functions that depend on it. The reactive functions configure outputs that update the tables, charts, and plots. When users change input values, the observeEvent object is triggered, which pushes the changes to a reactive object for configuration, and then updates the main dataset via mod_fun.R file. Finally, the main data in the reactiveValues object is altered, followed by a series of updating processes that cascade modifications through all outputs.

**Figure 1.**
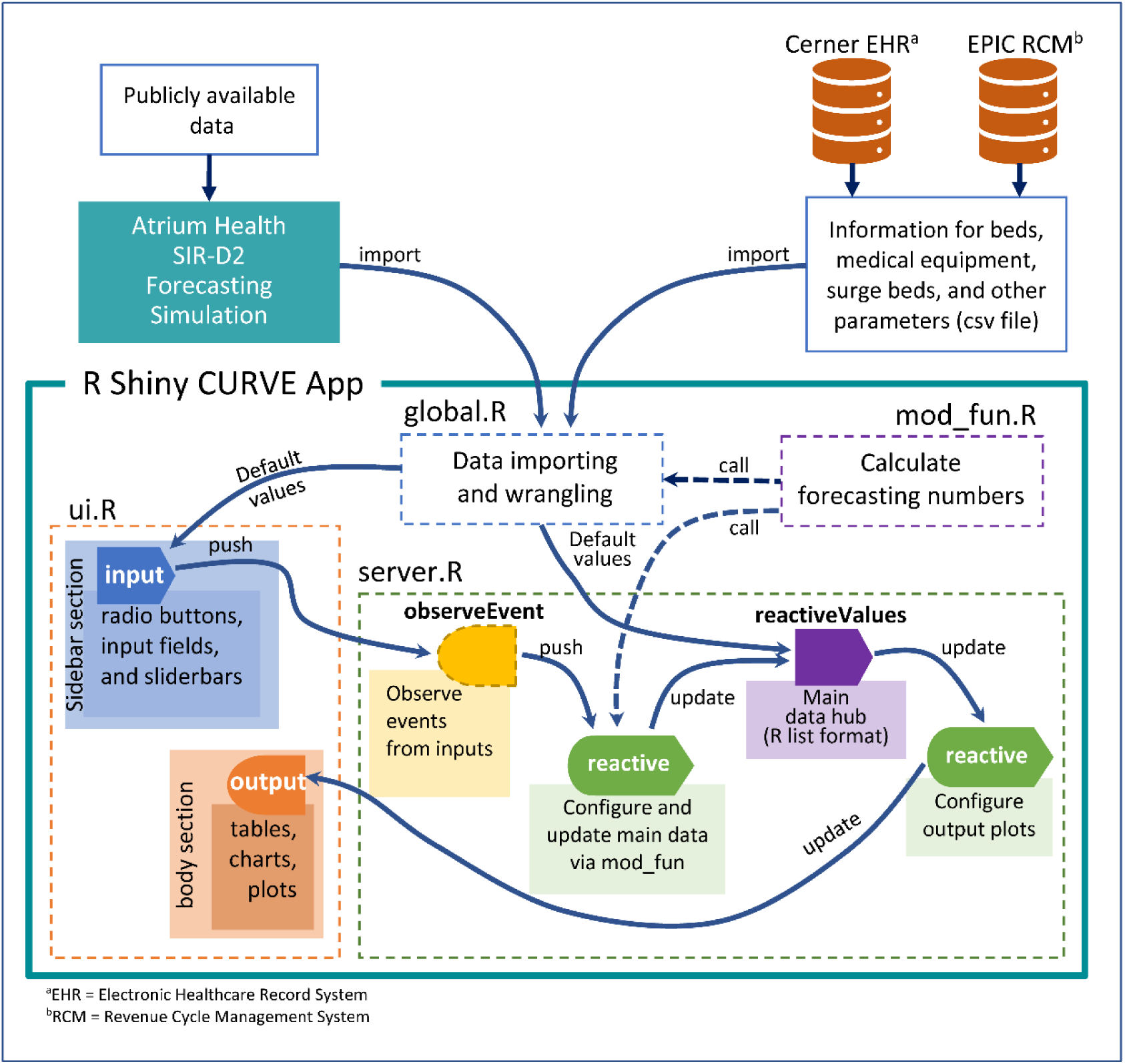
Schematic of the proposed CURVE app R Shiny reactivity diagram

The CURVE app provides health care leaders with: 1) predicted and observed COVID-19 hospital census by day for non-ICU beds, ICU beds, and ventilator utilization; 2) predicted and observed hospital admissions broken down by COVID-19 and non-COVID-19; 3) variance of the modeled prediction from actual occurrences; and 4) projections of the effect of decreasing social distancing measures on different dates. For planning purposes, users can choose from three preprogramed scenarios that depict a best-, moderate-, and worst-case scenario, or customize these configurations (Figure 2).

**Figure 2.**
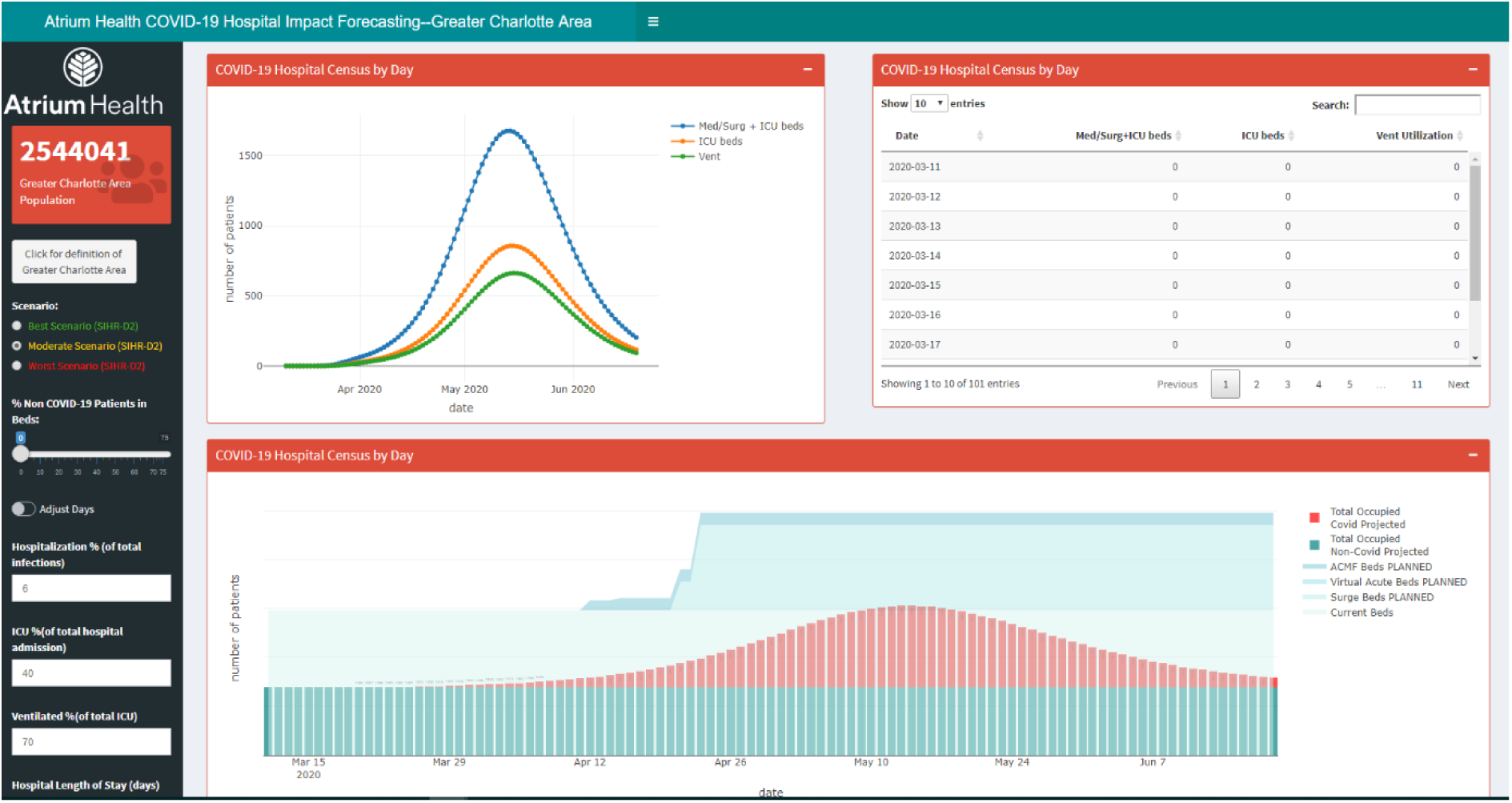
The output of the CURVE app.

**Figure 3.**
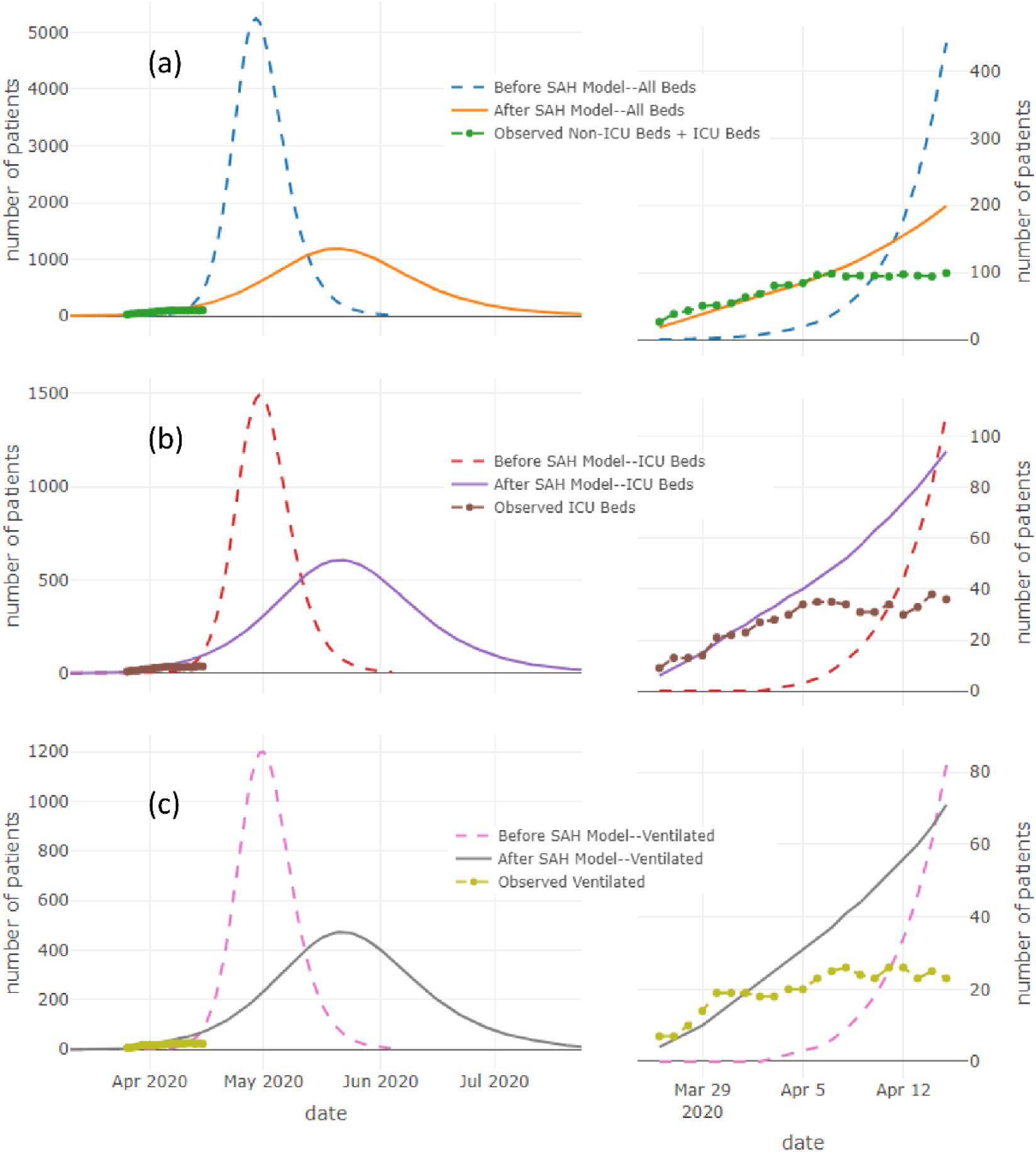
Model comparison in (a) all beds, (b) ICU beds, and (c) on ventilators between Before-SAH-Order and After-SAH-Order models. All graphs on right hand side are zoomed in results between Mar 26 and Apr 15, 2020.

In the model comparisons shown in Table 2, the AH before-SAH-order model projected higher volumes of hospitalized COVID-19 patients and earlier date to reach the peak (April 29 vs. May 20, 2020). The AH after-SAH-order model estimated a maximum number of COVID-19 patients on May 20, 2020 and the lowest RMSE in total-bed, ICU-bed, and patients on ventilator. Note that the observed all-bed and ICU beds experience a flattening phenomenon after April 6, 2020, which likely reflected the social distance and stay-at-home orders issued on March 30, 2020. The AH after-SAH-order model accordingly also reflects a flatter curve compared with the before-SAH-order model.

**Table 2.** Model comparison in peak date, maximum number of patients, and root mean square error (RMSE) between observed numbers and projections for moderate scenario parameters.

| Model | Peak Date | Maximum hospital bed census | <sup>a</sup>RMSE Between Observed Values and Projections |  |  |
| --- | --- | --- | --- | --- | --- |
|  |  |  | Total Beds | ICU Beds | On Ventilator |
| AH Before-SAH-Order | April 29, 2020 | 5,243 | 108.1 | 27.4 | 21.0 |
| AH After-SAH-Order | May 20, 2020 | 1,184 | 38.8 | 25.4 | 20.3 |

<sup>a</sup>RMSE=Root Mean Square Error.

## Discussion

We present a novel decision support app that incorporates locally calibrated forecasts with health system specific data and provides healthcare system leadership with timely actionable insights to guide decision making during the COVID-19 pandemic. The CURVE app provides information based upon local infectious patterns and allows for frequent updates as new forecast or health system data becomes available. By predicting utilization in the context of up-to-date local resources, such as hospital beds, ICU beds, and number of ventilators, it also provides a critical monitoring function (e.g. alignment of model predicting and observed bed statistics). Furthermore, health system leaders can adjust variables in the CURVE app to further explore different scenarios. Because this pandemic trajectory is expected to be highly depending on public policy and community behaviors, we present best-, moderate-, and worst-case scenarios, so that leaders can compare side-by-side the range of possible impacts on hospital plans. The reality of this variability can be seen as the observed hospitalizations for COVID-19 positive patients significantly flattened after April 6, 2020, and the trajectory of hospital census shifted from the moderate- to best-case scenario according to Table 3, Table 4, and Figure 4.

**Table 3.** Three scenarios of AH After-SAH-Order model comparison in peak date, maximum number of patients, and root mean square error between observed numbers and projections.

| Model | Peak Date | Maximum Hospital Bed Census | <sup>a</sup>RMSE Between Observed Values and Projections |  |  |
| --- | --- | --- | --- | --- | --- |
|  |  |  | Total Beds | ICU Beds | On Ventilator |
| Best-Case Scenario | May 20, 2020 | 584 | 33.1 | 9.6 | 8.4 |
| Moderate-Case Scenario | May 20, 2020 | 1,184 | 38.8 | 25.4 | 20.3 |
| Worst-Case Scenario | May 20, 2020 | 2,016 | 108.0 | 60.0 | 45.2 |

<sup>a</sup>RMSE=Root Mean Square Error.

**Table 4.** RMSE results applying the After-SAH-Order model before and after April 6, 2020 using the moderate-case and best-case scenario settings.

|  | <sup>a</sup>RMSE Between Observed Values and Projections |  |  |  |  |  |
| --- | --- | --- | --- | --- | --- | --- |
|  | Total Beds |  | ICU Beds |  | On Ventilator |  |
|  | Before Apr-6 | After Apr-6 | Before Apr-6 | After Apr-6 | Before Apr-6 | After Apr-6 |
| Moderate-Case Scenario | 7.8 | 58.6 | 4.5 | 38.4 | 6.1 | 30.3 |
| Best-Case Scenario | 36.2 | 28.4 | 11.0 | 7.4 | 9.0 | 7.5 |

**Figure 4.**
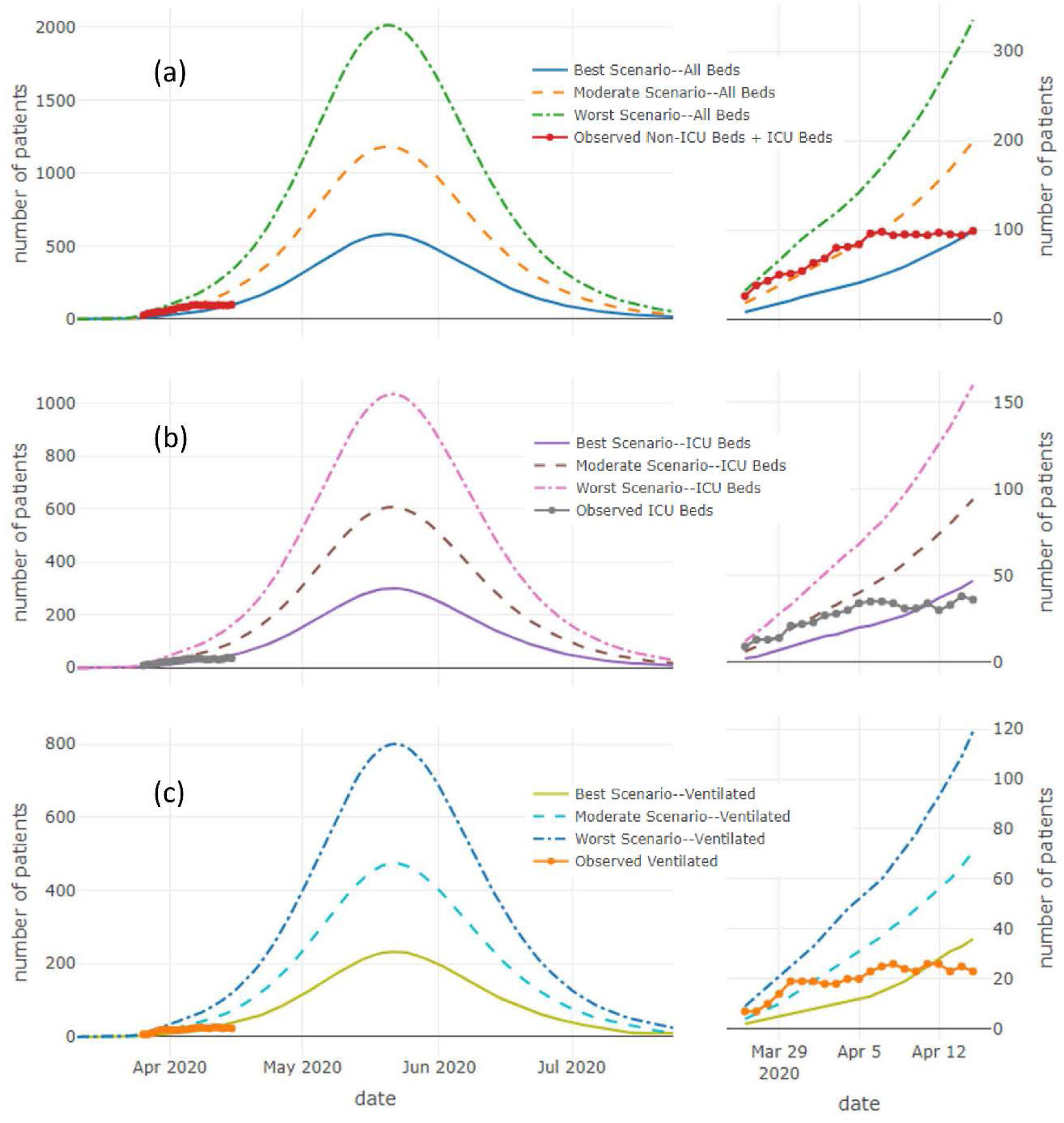
Model comparison in (a) all beds, (b) ICU beds, and (c) on ventilators between three models with best-case scenario, moderate-case scenario, and worst-case scenario settings, respectively. All graphs on right hand side are zoomed in results between Mar 26 and Apr 15, 2020.

In some cases, the user will have no idea as to a plausible value for the hospitalization proportion. One approach to determine a reasonable value is to plot the actual hospital census counts versus the date and modulate the hospital proportion until the predicted hospital census curve is parallel to the trend in the actual counts. Then add an offset to the predictions so that the curve provides a good fit to the actual counts. An offset that minimizes the RMSE is the difference between mean actual counts and the mean predictions. For a more refined approach, it is simple to write a routine in R that iterates through a sequence of possible hospitalization proportions, their corresponding offsets, and then select the hospital proportion that gives the smallest RMSE. In our case, a hospitalization proportion of 2.19% (with an offset of 43.6) gave the optimized fit, i.e., new RMSE=11.5. A more nuanced approach as the pandemic progresses would not assume one overall hospital proportion and use local linear trends.

Importantly, the pandemic behavior has evolved such that deterministic models or models not built to be responsive to local data will have poor fit with observed hospitalizations. Beyond providing a user-friendly and adaptable platform for hospital resource forecasting, the CURVE app also addresses a critical gap for health systems to have locally relevant, updated information. Other models and apps provided guidance to predict surge timing and health system demand requirements due to COVID-19; however, these are mostly built on national- and state-level data, are not updated frequently, and do not have actual trends of health system data that most accurately reflect local reality. ^6,8,19^ Our CURVE app addresses these shortcomings by relying upon modeling built on local data that is updated three times a week based upon observed infection and utilization rates. It is likely that more advanced models (e.g. Seasonal Autoregressive Integrated Moving Average (SARIMA), or Bayesian generalized additive model) will be required as the pandemic behavior continues to evolve. The CURVE app allows for these more advanced models to easily be consumed and available to leaders.

The CURVE app indicated that the after-SAH-order model with moderate-scenario parameter settings had good fit before April 6, 2020, and in near-real-time allowed our health system to see significant flattening of observed hospitalization for COVID-19 after April 6, 2020. This local response to social distancing, while immediately apparent in the CURVE app, would not be available to health system leaders relying on models that are neither built on local data nor with frequent recalibration to local trends. Whereas, in the CURVE app, leaders are able to immediately gain these ‘on the ground,’ local insights – indeed, in this case, that means shifting from a tracking on the moderate- to the best-case scenario.

Another benefit of the CURVE app is that it also accounts for non-COVID-19 healthcare utilization, so leaders have a full view of the health system’s capacity. While healthcare utilization for non-COVID-19 illness declined early in the pandemic due to canceling of all in-person elective care,^25^ this demand is also expected to surge back over time and increasingly become a contributor to overall health system resource consumption.^26–28^

There are some limitations and scope conditions that should be noted. First, although the CURVE app was developed for healthcare systems to insert local data using either csv files or through direct connection to their enterprise data warehouse, users still require R and R Shiny proficiency to modify or to add more tables, charts, or graphs. Second, in order to provide reliable, locally relevant forecasts, users must have access to local data and modeling expertise with frequent recalibration of the model to local context. If such changes are not monitored and accounted for, projections could be taken out of context, leading to erroneous conclusions.

## Conclusion

We developed an interactive hospital app using R Shiny to provides locally relevant, dynamic, timely information to guide health system decision making and pandemic preparedness. App frameworks like this will form the basis for data-informed health systems to be one step ahead of the current and future pandemics, so they are best prepared to serve their communities.

## Data Availability

Data not available due to legal restrictions

